# Respiratory Syncytial Virus infections in immunosuppressed adults <65 years old: a comparative analysis with Influenza and SARS-CoV2

**DOI:** 10.64898/2026.09.22.26363680

**Authors:** Maria Danysz, Eleanor Barnes, Sean H Lim, Alex G Richter, Beth Stuart, Michelle Willicombe

**Affiliations:** Department of Immunology and Inflammation, Centre for Inflammatory Disease, Imperial College London, London, UK; Nuffield Department of Medicine and Oxford NIHR Biomedical Research Centre, University of Oxford, Oxford, UK; Centre for Cancer Immunology, Faculty of Medicine, University of Southampton, UK; University Hospital Southampton NHS Foundation Trust, Southampton, UK; Clinical Immunology Service, School of Infection, Inflammation and Immunology, College of Medicine and Health, University of Birmingham, Birmingham, UK; Pragmatic Clinical Trials Unit Queen Mary University of London, London, UK; Hammersmith Hospital, Imperial College Healthcare NHS Trust, London, UK

**Author notes:** Contact Information: Dr Michelle Willicombe, Imperial College London, 9^th^ Floor, Commonwealth Building, Hammersmith Hospital Campus, London, UK, W12 0NN.

## Abstract

**Objectives:** Respiratory Syncytial Virus (RSV) vaccine policies have varied globally since their approval in 2023. In the UK, policy has adapted in line with National evidence generation such that from Autumn 2026 at-risk persons ≥65 years old will become eligible for vaccination. Immunosuppressed persons <65 years will still not be offered protection. The objective of this study is to investigate the relative burden of RSV infection by age and immunosuppressed status (IC).

**Design:** Retrospective observational cohort study using routine healthcare data from North West London Integrated Care System.

**Participants:** Patients with a confirmed diagnosis of a respiratory viral infection (RSV, Influenza A and B, SARS-CoV2) between October 2021 and July 2025.

**Main outcomes measures:** Prevalence and severity of RSV infection in immunosuppressed persons <65 years old, and to provide a comparative analysis with Influenza and SARS-CoV2 in a sub-cohort tested using a 4-in-1 combined rapid PCR test.

**Results:** During the study period, 1226 patients were diagnosed with RSV of whom 596 (48.6%) were <65 years old. In 1091 patients with clinical data, 341 (31.3%) were IC. Fifty (9.9%) patients <65 years old had severe infection (ICU admission or death within 28 days of diagnosis), of whom 14 (28.0%) were IC. Proportionally, 16.7% of all RSV-associated deaths and 55.1% of all ICU RSV-admissions occurred in adult patients <65 years old.

RSV accounted for 665 (10.1%) infections confirmed by 4-in-1 viral PCR testing. Eight-nine (13.4%), 172 (9.3%) and 448 (12.0%) patients with RSV, Influenza A and SARS-COV2 experienced severe infection, of whom 32 (36.0%), 82 (47.7%) and 156 (34.8%) were <65 years old respectively. Eight (25.0%), 20 (24.4%) and 52 (33.3%) of patients <65 years old with severe infection were IC. On adjusted analysis, having an immunosuppressive condition increased the odds of a severe SARS-CoV2 infection, OR (95% CI) 1.36 (1.08-1.71) but not a severe RSV or Influenza infection.

**Conclusions:** Severe RSV infections occur in persons <65 years old, with a significant proportion of patients having an immunosuppressive condition. Urgent research is needed to investigate the potential benefit of extending the current UK RSV vaccine policy to protect younger immunosuppressed adults.

## Introduction

The incidence, healthcare resource burden and outcome of Respiratory syncytial virus (RSV) infections have been underestimated in adults over time(1). Following regulatory approval of RSV vaccines in 2023 there is now a clear rationale to provide robust evidence on populations at-risk of infection who may benefit from protection afforded by vaccination. Surveillance programmes such as the Hospital-based Acute Respiratory Sentinel Surveillance (HARISS) in the UK have provided much needed data supporting RSV vaccine policy since their clinical implementation in 2024(2). The Joint Committee on Vaccination and Immunisation (JCVI) in the UK have been reactive to data generated, and as such, from September 2026, RSV vaccination scheduling will extend from persons ≥75 years, to include adults aged between 65-74 years old with chronic respiratory disorders or who are immunosuppressed(3). Immunosuppressed adults <65 years old will remain ineligible for vaccination in the UK(4).

Global RSV vaccination policies vary but most countries initially recommended vaccinating persons aged >60 years with co-morbidities and immunosuppressive conditions(5, 6). This age barrier has since been lifted with broad regulatory approval for vaccine use in at risk adults aged ≥18 years(7). If we are to learn from the COVID-19 pandemic, protecting immunosuppressed persons against infection is of paramount importance as although they compromise a small percentage of the population, infection not only portends a poor prognosis for an individual but may also result in higher healthcare resource needs and may pose a public health risk related to impaired viral clearance(8). From data available, immunosuppression or immunosuppressive conditions have been shown to be risk factors for severe RSV infection across all age groups(9-14). We also now have evidence that although RSV vaccine effectiveness is lower in immunosuppressed populations, vaccination still significantly reduces odds of severe infection(15-18). The question remains therefore, should younger immunosuppressed adults in the UK be offered vaccination against RSV too? Historical modelling data from the UK has suggested RSV associated deaths occur in adults <65 years, however granular data on the risk factors associated with these premature deaths are not known; an association with immunosuppression or other co-morbidities may be hypothesised(19).

Given the urgent need for preliminary evidence of the potential benefit of interventions against RSV infections in immunosuppressed persons, the aim of this study is to describe the relative prevalence and outcome of RSV infections by age and presence of an immunosuppressive condition. The secondary aim is to describe the frequency and outcome compared with SARS-CoV-2 and Influenza infections, vaccine preventable infections against which, younger immunosuppressed adults are vaccinated.

## Methods

The study uses the Imperial College Healthcare NHS Trust (ICHNT) - Clinical, Analytics, Research and Evaluation Informatics Environment (iCARE) Automated Data Model which contains data from patients who have had an inpatient, outpatient, or Accident and Emergency encounter at Imperial College Healthcare NHS Trust after 1^st^ January 2015. The use of the routinely collected data held within the iCARE system was approved by the South West – Central Bristol Research Ethics Committee (Health Research Authority reference: 21/SW/0120).

Respiratory syncytial virus infections were identified between 1^st^ October 2021 and 31^st^ May 2025 from either positive tests reported from North West London Pathology from patients tested at one of the 5 ICHNT hospitals, or from coding from a hospital episode alone. Diagnoses related to the RSV hospital episode included one of the following codes: B97.4, J12.1, J21.0, J20.5 (ICD-10); 6415009, 55735004, 195739001, 57089007, 195881003, 79479005 (SNOMED CT UK).

The RSV testing platforms included either the combined rapid polymerase chain reaction (PCR) test for Influenza A/B, RSV and SARS-CoV-2, RSV testing as part of a respiratory virus PCR screen, or as part of a pneumonia PCR panel. For positive tests, the date and time of sample collection was used as the diagnosis date. For code diagnoses, the start date and time of the episode during which an RSV diagnosis was recorded was used as the diagnosis date and time. Positive tests or diagnoses recorded more than 60 days after a previous positive test or diagnosis and not during the same hospital stay were considered as new infections. The first positive test or diagnosis after 1^st^ October 2021 was always considered as a new infection. A new infection season was counted from the 1^st^ day in October each year; with winter running from 1^st^ October to 31^st^ March, and summer running from 1^st^ April to 30^th^ September.

For the comparative analysis of Influenza A/B, RSV and SARS-CoV-2, only data from testing performed by the combined rapid PCR test between 1^st^ October 2021 and 13^th^ July 2025 was utilised. Where patients were diagnosed with ≥1 virus type during the study period, infections were considered independently. Repeated positive tests of the same virus were considered re-infections if sampled 30 days, 60 days, and 90 days after previous infection for influenza, RSV, and SARS-CoV-2 respectively.

Only patients ≥18 years old were included in this analysis.

### Outcomes

An RSV-associated hospital admission was recorded as an admission episode occurring concurrently or within 28 days of a positive swab. Severe infection was defined as infection followed by admission to an intensive care or high dependency unit, or death within 28 days. Where a patient did not die in hospital, only the month and year of death were available. For those patients, the 15^th^ of the month was assumed as the date of death.

Risk factors for severe infection were defined as having an ICD-10 code associated with immunosuppression recorded during an inpatient stay up to a year before a new infection, or having an immunosuppression-associated condition recorded as a long-term problem at any point before their infection (*Supplementary Information*, **Appendix 1**). Immunosuppressive conditions used included those listed by The Green Book, the Immunisation against infectious Disease guideline in the UK(20). The conditions include(20): immunosuppression due to disease or treatment, including patients undergoing chemotherapy, patients undergoing radical radiotherapy, solid organ transplant recipients, bone marrow or stem cell transplant recipients, HIV infection at all stages, multiple myeloma or genetic disorders affecting the immune system. Individuals who are receiving immunosuppressive or immunomodulating biological therapy including individuals treated with or likely to be treated with systemic steroids for more than a month at a dose equivalent to prednisolone at 20mg or more per day for adults. Anyone with a history of haematological malignancy, and those who require long term immunosuppressive treatment for conditions including, but not limited to, systemic lupus erythematosus, rheumatoid arthritis, inflammatory bowel disease, scleroderma and psoriasis.

### Statistical Analysis

Odds ratios of ICU admission, death, or composite severe outcome were estimated using logistic regression. Only patients with hospital diagnosis data recorded up to a year before their viral infection were included in the models. Where one patient had repeat infections during the study period, only the first infection recorded during that time was used in the model. Models were adjusted for age and sex. Age groups were categorised to reflect RSV vaccination policy. Ethnicity was categorised as follows: White (White British, White Irish, any other White background), Asian (Bangladeshi, Indian, Pakistani, Chinese, any other Asian background), Black (African, Caribbean, any other Black background), Other (any Mixed background, any other ethnic group, other – unknown or unstated). Proportions were compared with chi-square tests.

Statistical analyses were performed using R Notebooks in Azure Machine Learning.

## Results

Between 1^st^ October 2021 and 31 May 2025, RSV infection was recorded 2973 times in 1226 adults aged >18 years old. After removing diagnoses recorded within 60 days of the first episode, there were 1250 new infections throughout the entire period; 1203 patients had only one infection, 22 patients had two infections, and one patient had an infection three times. The clinical demographics of patients diagnosed with RSV infection are outlined in **Table 1**. Patients were aged between 18 and 104, with 596 (48.6%) patients <65 years old. The majority were female, white, and living in deprived areas. Of the 1226 patients, 914 (74.6%) had an RSV infection episode associated with a hospital admission, of which, 402 (44.0%) occurred in patients <65 years. The median length of stay was 5 (IQR 1-16) days. Most infections, 1114/1226 (90.9%), were diagnosed during a winter season.

**Table 1.**
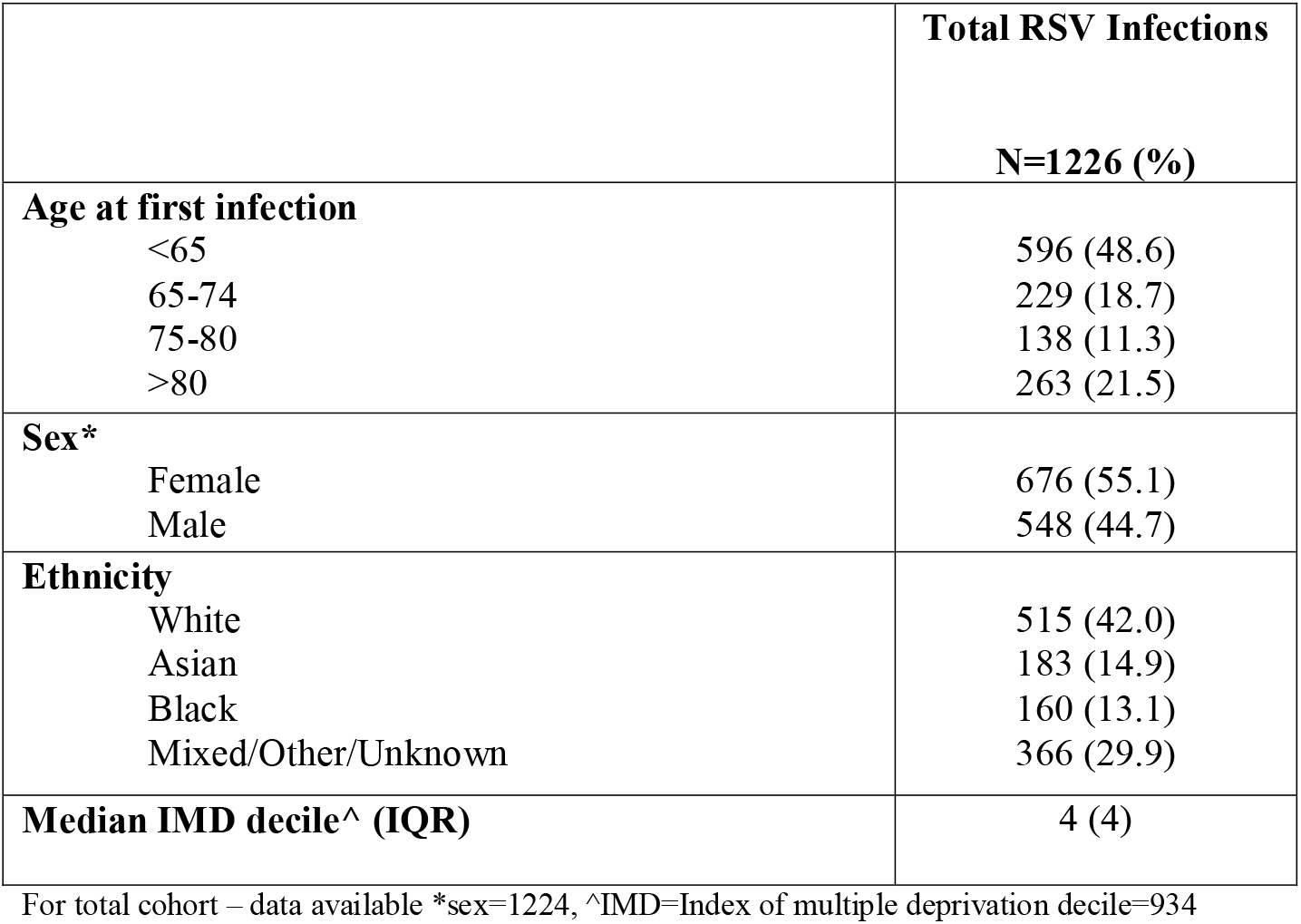
Clinical demographics and outcomes of patients diagnosed with Respiratory Syncytial Virus infection.

Data on clinical co-morbidity was available for 1091 patients, of these, 341(31.3%) had ≥1 immunosuppressive condition, **Table 2**. The proportion of RSV infected patients <65 years old and >65 years old with an immunosuppressive condition was 188 (37.4%) and 153 (26.0%) respectively, p<0.01 (*Supplemental Information*, **Table S1)**. Of the 1091 patients, 903 (83.7%) had an RSV infection episode associated with a hospital admission, of which, 398 (44.1%) occurred in patients <65 years. The proportion of <65 year and >65 year olds with ≥1 immunosuppressive condition with an RSV associated hospital admission was 161/398 (40.5%) and 135/505 (26.7%) respectively, p<0.01. Specialty of admitting team and median age of patients during the admission where infection was diagnosed is shown in *Supplemental Information*, **Table S2**. The distribution of cases by year, covering winter 2024-2025 when the RSV vaccine was introduced is shown in *Supplemental Information*, **Figure S1**. In the year prior to RSV vaccine roll out, 2023-2024, 63/435 (14.4%) of RSV cases were diagnosed in persons aged between 75-79 years old compared with 28/390 (7.2%) in 2024-2025, p<0.01, with 5 reinfections.

**Table 2.** Clinical characteristics and outcomes of patients diagnosed with Respiratory Syncytial Virus infection with co-morbidity data available.

|  | **Total RSV Infections** | **Severe RSV Infection** | **RSV ICU Admission** | **RSV Death** |
| --- | --- | --- | --- | --- |
|  | **N=1091 (%)** | **N= 141 (%)** | **N=78 (%)** | **N=78 (%)** |
| **Age at first infection** |  |  |  |  |
| <65 | 503 (46.1) | 50 (35.5) | 43 (55.1) | 13 (16.7) |
| 65-74 | 207 (19.0) | 27 (19.1) | 16 (20.5) | 13 (16.7) |
| 75-80 | 132 (12.1) | 19 (13.5) | 11 (14.1) | 12 (15.4) |
| >80 | 249 (22.8) | 45 (31.9) | 8 (10.3) | 40 (51.3) |
| **Sex\*** |  |  |  |  |
| Female | 600 (55.0) | 72 (51.1) | 38 (48.7) | 40 (51.3) |
| Male | 491 (45.0) | 69 (48.9) | 40 (51.3) | 38 (48.7) |
| **Ethnicity** |  |  |  |  |
| White | 460 (42.2) | 65 (46.1) | 33 (42.3) | 41 (52.6) |
| Asian | 169 (15.5) | 14 (14.9) | 13 (16.7) | 8 (10.3) |
| Black | 150 (13.7) | 19 (13.5) | 12 (15.4) | 9 (11.5) |
| Mixed/Other/Unknown | 312 (28.6) | 36 (25.5) | 20 (25.6) | 20 (25.6) |
| **Median IMD decile^ (IQR)** | 4 (4) | 3(4) | 4 (4) | 3 (4) |
| **≥1 Immunosuppressive condition** | 341 (31.3) | 32 (22.7) | 18 (23.1) | 20 (25.6) |

Of the 1091 patients with co-morbidity data, 141 (11.5%) had a severe RSV infection; 78 (6.4%) patients were admitted to intensive care within 28 days of infection and 78 (6.4%) died within 28 days of infection. The median length of stay in intensive care was 6 (IQR 9.75) days. Clinical characteristics of patients with severe infection are shown in **Table 2**. The proportion of patients with severe infection who had an immunosuppressive condition was 14 (28.0%) and 18 (19.9%) in patients <65 and ≥65 years old respectively, p=0.27 (*Supplemental Information*, **Table S1)**. The specialty of the team and the median age of the patients during the severe infection admission is shown in Supplemental *Information*, **Table S3**.

Forty-one (55.1%) infected patients admitted to ICU were <65 years old, whilst 65 (83.3%) patients who died were ≥65 years old. On unadjusted analysis, the presence of an immunosuppressive condition was associated with reduced odds of a RSV associated severe infection, OR 0.61 (95% CI 0.4-0.92), whilst older age was associated with increased odds of severe infection, OR 1.22 (95% CI 1.06-1.41). There was no association between female sex and severe outcome, OR 0.81 (95% CI 0.56-1.15). After adjusting for age and sex, the presence of an immunosuppressive condition was no longer associated with odds of severe infection, OR 0.68 (95% CI 0.43-1.02).

For the comparative analysis between Influenza A/B, RSV and SARS-CoV2, data from 245,383 tests performed in 39,400 patients who were swabbed using the combined 4-in-1 PCR test was studied. The clinical characteristics of patients who were swabbed are shown in *Supplemental Information*, **Table S4**. In total, 9326 (23.7%) individual patients tested positive for at least one virus. The number of new infections by year and by season is shown in *Supplementary Information*, **Figure S2** and **S3** respectively. Influenza A became the dominant viral infection in the year 2024-2025, exceeding the frequency of SARS-CoV2 infections for the first time during the period of data collection, **Figure 2**. Differing from Influenza and RSV, SARS-CoV2 infections occurred both in winter and summer months (*Supplementary Information*, **Figure S3)**.

The clinical characteristics of patients testing positive for Influenza A/B, RSV or SARS-CoV2 on the combined PCR test is shown in *Supplementary Information*, **Table S5**. Co-morbidity data was available in 6070 individual patients testing positive for 6574 infections, with 1512/6574 (23.0%) infections occurring in patients having ≥1 immunosuppressive condition, **Table 3**. Of the 462/6070 (7.6%) patients who were diagnosed with ≥1 infection type during the study period, 235 (50.9%) were <65 years old and 90 (19.5%) were immunosuppressed.

**Table 3.** Clinical characteristics of 6070 patients testing positive for 6574 infections Influenza, RSV or SARS-CoV-2 with co-morbidity data.

|  | **Influenza A**<br>**N= 1850 (%)** | **Influenza B**<br>**N= 320 (%)** | **RSV**<br>**N= 665(%)** | **SARS-CoV2**<br>**N= 3739 (%)** |
| --- | --- | --- | --- | --- |
| **Sex** |  |  |  |  |
| Female | 1042 (56.3) | 206 (64.4) | 381 (57.3) | 1966 (52.6) |
| Male | 808 (43.7) | 114 (35.6) | 284 (42.7) | 1773 (47.4) |
| **Age** |  |  |  |  |
| <65 | 949 (51.3) | 273 (85.3) | 273 (41.1) | 1503 (40.2) |
| 65-74 | 308 (16.6) | 15 (4.7) | 135 (20.3) | 700 (18.7) |
| 75-80 | 212 (11.5) | 15 (44.7) | 87 (13.1) | 503 (13.5) |
| 80+ | 381 (20.6) | 17 (5.3) | 170 (25.6) | 1033 (27.6) |
| **Ethnicity** |  |  |  |  |
| White | 726 (39.2) | 87 (27.2) | 302 (45.4) | 1741 (46.6) |
| Asian | 279 (15.1) | 66 (20.6) | 100 (15.0) | 450 (12.0) |
| Black | 274 (14.8) | 61 (19.1) | 85 (12.8) | 517 (13.8) |
| Mixed/Other/Unknown | 571 (30.9) | 106 (33.1) | 178 (26.8) | 1031 (27.6) |
| **IMD decile IQR)\*** | 4 (3) | 4 (3) | 4 (4) | 4 (3) |
| **≥1 Immunosuppressive condition<sup>^</sup>** | 354 (19.1) | 62 (19.4) | 188 (28.2) | 908 (24.3) |

Seven hundred and twenty-six (11.0%) of 6574 infections were severe. The clinical characteristics of patients with severe Influenza A, RSV or SARS-CoV2 are shown in **Table 4**. Only 17 (5.3%) Influenza B infections resulted in admission to ICU or death within 28 days and were not analysed further. The proportion of patients with severe Influenza A, RSV or SARS-CoV2 who were <65 years old was 82 (47.7%), 32 (36.0%) and 156 (34.8%) respectively, **Table 4**. Of the patients <65 years old with severe infection, 20 (24.4%) with Influenza A, 8 (25.0%) with RSV and 52 (33.3%) with SARS-CoV2 had an immunosuppressive condition (Supplementary *Information*, **Table S6)**. On adjusted analysis, having ≥1 immunosuppressive condition increased the odds of severe SARS-CoV2 infection, OR 1.36 (95% CI 1.08-1.71) but not a severe Influenza or RSV infection, **Table 5**.

**Table 4.**
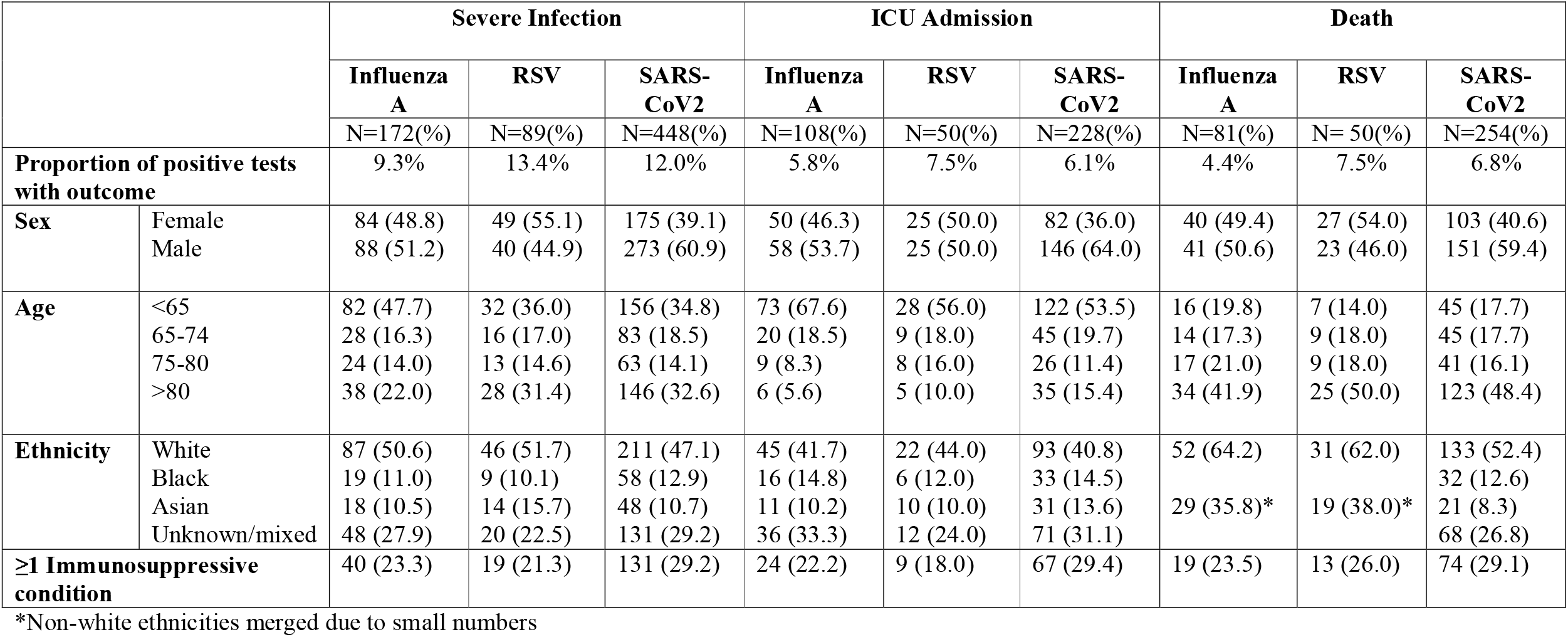
Clinical characteristics of 709 patients with severe Influenza A, RSV or SARS-CoV-2 infection requiring intensive care admission and/or dying within 28 days of infection.

**Table 5.** Odds of severe infection, ICU admission and death in patients with ≥1 immunosuppressive condition testing positive for Influenza-A, RSV or SARS-CoV2.

|  | Severe Infection |  | ICU Admission |  | Death |  |
| --- | --- | --- | --- | --- | --- | --- |
|  | OR (95% CI) | P value | OR (95% CI) | P value | OR (95% CI) | P value |
| **Influenza-A** |  |  |  |  |  |  |
| Unadjusted | 1.28 (0.86-1.85) | 0.21 | 1.25 (0.77-1.97) | 0.35 | 1.20 (0.67-2.03) | 0.52 |
| Adjusted^ | 1.30 (0.87-1.89) | 0.19 | 1.12 (0.68-1.77) | 0.65 | 1.51 (0.83-2.60) | 0.16 |
| **RSV** |  |  |  |  |  |  |
| Unadjusted | 0.62 (0.35-1.05) | 0.09 | 0.54 (0.24-1.08) | 0.10 | 0.82 (0.40-1.56) | 0.56 |
| Adjusted^ | 0.66 (0.37-1.12) | 0.14 | 0.46 (0.20-0.93) | 0.04 | 1.18 (0.56-2.34) | 0.65 |
| **SARS-CoV2** |  |  |  |  |  |  |
| Unadjusted | 1.29 (1.03-1.61) | 0.03 | 1.24 (0.91-1.68) | 0.16 | 1.30 (0.97-1.73) | 0.07 |
| Adjusted^ | 1.36 (1.08-1.71) | 0.01 | 1.11 (0.81-1.51) | 0.52 | 1.63 (1.47-1.83) | 0.001 |

^adjusted for age and sex

In total, 386/6254 (6.2%) Influenza A, RSV or SARS-CoV2 infections led to an ICU admission. The majority of ICU admissions occurred in patients <65 years; 73 (67.6%), 28 (56.0%) and 122 (53.5%) in Influenza A, RSV and SARS-CoV2 infections respectively. The proportion of <65-year-olds admitted to ICU who had an immunosuppressive condition was 16 (22.0%) and 33 (27.0%) for Influenza A and SARS-CoV2 respectively, with <5 patients with RSV infection being immunosuppressed (*Supplemental Information*, **Table S6)**. On unadjusted analysis, having ≥1 immunosuppressive condition did not influence the odds of requiring an ICU admission in the context of any infection, however it was associated with reduced odds in RSV infected patients on adjusted analysis, OR 0.46 (95% CI 0.20-0.93), **Table 5**.

Overall, 385/6254 (6.2%) patients testing positive for either Influenza A, RSV or SARS-CoV-2 died within 28 days of diagnosis. Of the deaths, 16 (19.8%), 7 (14.0%) and 45 (17.7%) occurred in patients <65 years old with Influenza A, RSV and SARS-CoV2 infection respectively. The proportion of <65-year-olds who died who had an immunosuppressive condition was 6 (37.5%) and 22 (48.9%) for Influenza A and SARS-CoV-2 respectively, with <5 patients with RSV infection being immunosuppressed (*Supplemental Information*, **Table S6**). Having ≥1 immunosuppressive condition in the context of any infection did not influence the odds of death on unadjusted analysis, however, odds of death were increased in immunosuppressed patients testing positive for SARS-CoV2 on adjusted analysis, OR 1.63 (95% CI 1.47-1.83), **Table 5**.

## Discussion

This study has shown that almost half of RSV infections are diagnosed in adults <65 years old and severe infections occur in up to 9.9% of these patients. This corresponds to upper estimates of 2.6% and 8.5% of patients <65 years old who were diagnosed with RSV either dying or admitted to ICU respectively. Proportionally from all RSV infections, 16.7% of RSV-associated deaths and 55.1% of all ICU RSV-admissions occurred in adult patients <65 years old. These data suggest RSV infections in patients <65 years are clinically significant and collectively may represent an important healthcare burden.

Within the limitations of the study, we were able to report that a significant proportion of patients <65 years old with RSV infection had an immunosuppressive condition. Although there was no denominator data for persons at risk, from prior estimates of the number of immunosuppressed persons <65 years old in the general population, we can confidently state that such persons appear to be over-represented in our RSV cases(21). Such a comment would also be supported from data available showing that immunocompromise is a risk factor for symptomatic RSV infection(11, 22). Whilst the inclusion of patients with clinical coding only in our analysis is likely to bias the association between immunosuppression and severity, the lower estimates of 27.8% of RSV+ patients being immunosuppressed still surpasses proportions estimated in the general population(21). It should also be considered that the true proportion of immunosuppressed adults in our study may be an underestimate, as diagnosis of underlying conditions reliant on clinic coding are often fraught with quality concerns, especially in complex multimorbid populations.

Independent of co-existing immunosuppressive conditions, understanding why other younger persons experienced severe infection will be important. One explanation may be the presence of other immune vulnerable co-morbidities not currently included in the definition of an immunosuppressive condition for vaccine policy(20). For example, almost 20% of patients with severe RSV were admitted under nephrology, and these patients had a median age of 59 years. Nephrology services at ICHNT primarily cares for patients with end stage kidney disease (dialysis and transplant), and whilst patients requiring dialysis are recognised to be at risk of infection, they are not officially classified as being immunosuppressed in the UK(23, 24). Data from the HARISS programme has highlighted the importance of co-morbidity as a risk factor for infection outcome in persons ≥65 years old, and the data presented herein supports the need for similar evidence in younger co-morbid patients(2).

Intensive care admissions and 30-day mortality rates associated with RSV infection in ≥65 year olds in the UK have been reported to be 1.2% and 10.6% respectively, compared with 5.6-6.0% and 10.3-11.1% in our cohort(2). Why we found higher estimates for ICU admissions in our cohort is unclear but warrants further exploration. Fortunately, the national statistics for RSV infections in older persons are now being transformed following the implementation of the RSV vaccines, with recent data showing a vaccine effectiveness (VE) against RSV-associated hospitalisation of 75 (69-80)% in 75-79 year olds in the first season of roll out(25). Multiple real-world studies globally have demonstrated similar levels of protection, mirroring the results of the vaccine efficacy studies(15). The pivotal studies for the 3 approved RSV vaccines, the AReSVi-006 Study (Arexy, GSK), RENOIR (Abrysvo, Pfizer) and ConquerRSV (mRESVIA, Moderna) collectively reported vaccine efficacies of between 66.7-82.6% against RSV-infection(26-28). Unfortunately, immunosuppressed persons were excluded from all these studies, and whilst subsequent immunobridging studies have since been performed in 18-59 year olds for both Arexy and Abrysvo in at-risk persons, these too excluded immunosuppressed persons(29, 30). An Arexy immunogenicity study has been conducted in solid organ transplant recipients which reports acceptable rates of humoral responses, however a peer reviewed publication is currently awaited(31). At present therefore, data on VE in immunosuppressed persons is reliant on real world data which is currently confined to persons ≥60 years. However, the data appears reassuring, as whilst effectiveness is reported to be less in immunocompromised older persons, it is still showing significant protection. Data from the UK in 75-79 year olds reports a VE against hospitalisation of 65 (49-77)% in immunocompromised persons versus 78 (71-84)% in the non-immunocomprised(25). Whilst a recent systematic review reports a comparative VE of 69 (66-73)% against RSV-associated hospitalisation in immunocompromised persons >60 years old(15).

A comparative analysis of outcomes of Influenza, RSV and SARS-CoV2 infections is of interest given that younger UK immunosuppressed adults are offered annual and biannual vaccination against Influenza and SARS-CoV2 respectively. Several studies have compared outcomes of these respiratory viral infections but to our knowledge none specifically in immunosuppressed populations. Two previous studies have captured persons testing positive for respiratory viral infections in the community, and suggested that SARS-CoV2 infection has remained the highest risk for a more severe disease course, but that risk appeared to be changing over time(32, 33). In another study of hospitalised adult patients, SARS-CoV2 infection appears to continue to have the worse prognosis, although this has been inconsistently reported and maybe more similar than previously assumed(34, 35). In this study, the number of RSV infections diagnosed was less than either SARS-CoV2 or Influenza A over the study period, but severity of infections when diagnosed were similar. However, consistent with extensive published SARS-CoV2 related evidence, this study showed that the adjusted odds of severe infection was increased for SARS-CoV2 infection in the presence of an immunosuppressive condition(23). In contrast, immunosuppression was not associated with adverse outcomes following RSV or influenza infection. Unexpectedly, we report that immunosuppression was associated with a reduced likelihood of an RSV-associated ICU admission, although importantly this was not accompanied by a reduction in mortality. This finding should be interpreted cautiously, ICU admission is influenced not only by infection severity but also by clinical decision-making, co-morbidity, frailty and ceilings of care, which may differ in immunosuppressed populations. In addition, immunosuppressed patients have greater healthcare contact and may undergo respiratory viral testing at a lower threshold, potentially increasing ascertainment of less severe RSV infections compared with the wider population. The broad definition of immunosuppression used in this study also encompasses clinically and biologically heterogeneous conditions that may confer different risks. These factors, together with the relatively small number of RSV-associated ICU admissions, may contribute to the observed association. The absence of a corresponding reduction in mortality provides no evidence that RSV infection is less clinically important in immunosuppressed patients and highlights the limitations of using ICU admission alone as a marker of disease severity in this population. In other studies of immunocompromised persons with respiratory infections excluding SARS-CoV2, viral pathogen does not appear to influence prognosis following ICU admission, which is more related to co-morbidity and clinical status; unfortunately, we did not have access to data related to clinical parameters(36, 37). Regardless of virus type, it is recognised that vaccine preventable infections are high in immunocompromised persons in all ages, and are associated with significant morbidity and mortality(38). This is the rationale for some countries endorsing RSV vaccination in immunosuppressed adults of all ages now the vaccines have regulatory approval to be used in persons ≥18 years old(7).

This study has significant limitations invariably associated with co-morbidity data reliant on clinical coding as already outlined. The reliability of viral infection diagnoses however was enhanced with incorporation of pathology data. Data on infection severity (ICU admissions and death) was also complete due to regulatory capture in surveillance programmes. Missing data was unlikely to be at random, which therefore introduces potential significant biases into our conclusions. However, the data presented highlights that the health care burden of RSV infections is not confined to persons ≥65 years, and supports the need to urgently generate further evidence on the potential benefit of RSV vaccination for younger adults.

To conclude, persons of all ages who are immunosuppressed are recognised to be at risk of severe RSV infection, and real-world data has shown that vaccination reduces risk of severe infections(17, 39). Consideration is now needed to offer the RSV vaccine to younger immunosuppressed adults in the UK. Unfortunately, it appears that barriers for evidence generation in immunocompromised persons remain unchanged from the COVID-19 pandemic; they are excluded from vaccine studies and real-world evidence is limited by their accurate identification in the UK. The HARISS surveillance is an excellent resource but only collects data on persons ≥65 years, surveillance programmes to capture data on all immunosuppressed populations of all ages is needed, not only for RSV but to inform optimal protection strategies for other respiratory viruses and future pandemic preparedness.

## Acknowledgments

This study was supported by the Imperial Clinical Analytics Research and Evaluation (iCARE) Secure Data Environment and used the iCARE and Whole System Integrated Care team and data resources. MD was funded by the Department of Health and Social Care (DHSC) via NIHR Grant code NIHR135830. EB acknowledges Oxford NIHR Biomedical research centre and is supported by the NIHR as a Senior Investigator. MW is supported by the National Institute for Health Research (NIHR) Biomedical Research Centre (BRC) based at Imperial College Healthcare NHS Trust and Imperial College London. SHL is support by a Cancer Research UK Advanced Clinician Scientist Award. The research was reviewed by representatives from the patient experience research centre at Imperial College London.

## Data Availability Statement

Data used in this study were held within the iCARE Secure Data Environment, and patient data are not available for sharing on account of privacy regulations. Code used for the study and aggregate data outputs are available from the authors upon reasonable request

