## Supplemental Information for "Respiratory Syncytial Virus infections in immunosuppressed adults <65 years old: a comparative analysis with Influenza and SARS-CoV2"

**Supplemental Table S1. Clinical outcome of patients 1091 patients diagnosed with Respiratory Syncytial Virus infection according to age and presence of immunosuppressive condition (IC)**

|  | | **Total RSV Infections**  **N=1091 (%)** | | **Severe RSV Infection**  **N=141(%)** | | **RSV ICU Admission**  **N=78 (%)** | | **RSV Death**  **N=78(%)** | |
| --- | --- | --- | --- | --- | --- | --- | --- | --- | --- |
|  |  | IC  N=341 (%) | Non-IC  N=750 (%) | IC  N=32 (%) | Non-IC  N= 109(%) | IC  N=18 (%) | Non-IC  N=60 (%) | IC  N=20 (%) | Non-IC  N= 58(%) |
| **Age (Years)** | **<65**  **≥65** | 188 (55.1)  153 (44.9) | 315 (42.0)  435 (58.0) | 14 (43.8)  18 (56.2) | 36 (33.0)  73 (67.0) | 10 (55.6)  8 (44.4) | 33 (55.0)  27 (45.0) | 7 (35.0)  13 (65.0) | 6 (10.3)  52 (89.7) |

**Supplemental Table S2. Speciality of caring team and median age of patients during the index admission where RSV was diagnosed**

| **Specialty** | **No. of patients** | **% of patients** | **Median Age (Yrs)** | **IQR** |
| --- | --- | --- | --- | --- |
| Clinical haematology | 126 | 23.60% | 58 | 2 |
| Geriatric medicine | 103 | 19.29% | 80 | 16 |
| General Internal Medicine | 98 | 18.35% | 73.5 | 25.75 |
| Respiratory medicine | 81 | 15.17% | 70 | 19.75 |
| Endocrinology | 72 | 13.48% | 75 | 19 |
| Gastroenterology | 55 | 10.30% | 73 | 19 |
| Acute internal medicine | 53 | 9.93% | 72 | 23 |
| Nephrology | 47 | 8.80% | 59 | 19 |
| Accident & Emergency | 34 | 6.37% | 74.5 | 24.25 |
| Cardiology | 32 | 5.99% | 70 | 19.75 |
| Obstetrics | 25 | 4.68% | 35 | 6 |
| Rheumatology | 20 | 3.75% | 71.5 | 14.25 |
| Medical oncology | 17 | 3.18% | 63 | 10 |
| Trauma & orthopaedics | 15 | 2.81% | 72 | 26 |
| Haematology | 13 | 2.43% | 63 | 15 |
| Critical care medicine | 11 | 2.06% | 65 | 26 |
| Clinical pharmacology | 10 | 1.87% | 71.5 | 15.5 |
| Neurology |  |  | 70 | 11.75 |
| General Surgery | 9 | 1.69% | 70 | 22 |
| Infectious diseases | 8 | 1.50% | 57.5 | 37.5 |
| ENT | 5 | 0.94% | - | - |
| Anaesthetics |  |  | - | - |
| Clinical oncology |  |  | - | - |
| Other* | <5 | <1% | - | - |

*Other: Genito-urinary medicine, neurosurgery, cardiothoracic surgery, rehabilitation, urology, gynaecology, paediatrics, plastic surgery, radiology

Note: patients under multiple specialties counted separately for each specialty.

IQR: Interquartile range; - not includes as <5 patients

**Supplemental Figure S1. Distribution of RSV diagnoses by age group by year from 1^st^ October 2021.**

In the year 2023-2024, 63/435 (14.4%) of cases were diagnosed in persons aged between 75-79 years compared with 28/390 (7.2%) in 2024-2025, p<0.01.

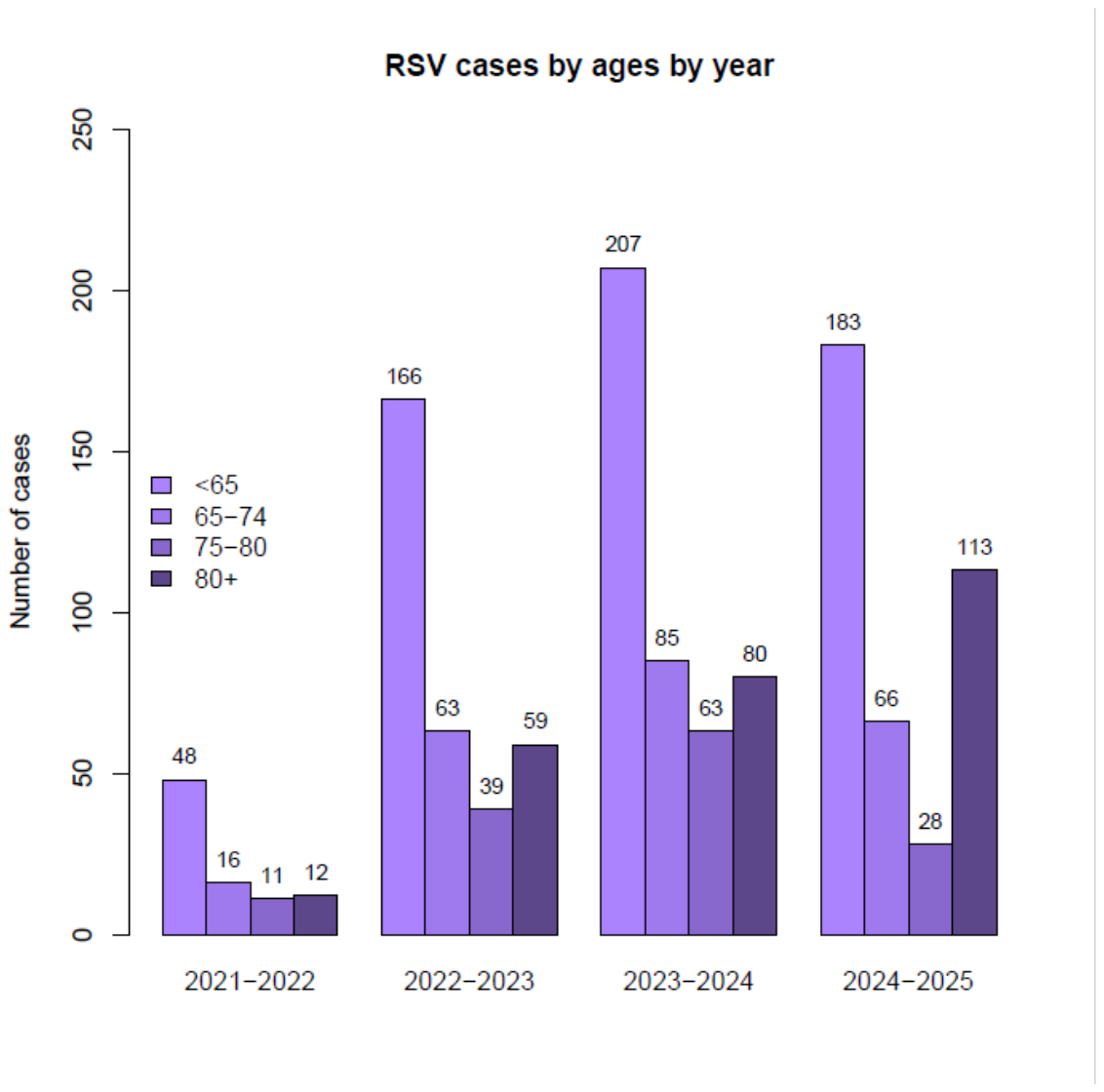

**Supplemental Table S3. Speciality of caring team and median age of patients during the admission where a severe RSV infection was diagnosed**

| **Specialty** | **No. of patients** | **% of patients** | **Median Age (Yrs)** | **IQR** |
| --- | --- | --- | --- | --- |
| Respiratory medicine | 18 | 23.4% | 75.5 | 16 |
| Geriatric medicine | 17 | 22.1% | 81 | 8 |
| Nephrology | 15 | 19.5% | 55 | 19 |
| Gastroenterology | 14 | 18.2% | 76.5 | 15.5 |
| General internal medicine | 14 | 18.2% | 80 | 13.25 |
| Endocrinology | 13 | 16.9% | 79 | 9 |
| Cardiology | 10 | 13.0% | 63 | 14.5 |
| Acute internal medicine | 7 | 9.1% | 85 | 2.5 |
| Clinical haematology |  |  | 60 | 5 |
| Other* | <5 | <6.5% | - | - |

*Other: Anaesthetics, critical care medicine, accident & emergency, general surgery, trauma & orthopaedics, rheumatology, neurology, clinical pharmacology, cardiothoracic surgery, neurosurgery, plastic surgery, medical oncology, infectious diseases, genito-urinary medicine

Note: patients under multiple specialties counted separately for each specialty.

**Supplemental Table S4. Clinical characteristics of patients being tested with the combined Influenza, RSV and SARS-CoV-2 assay**

|  | **N=39400 (%)** |
| --- | --- |
| **Sex** |  |
| Female  Male | 20764 (52.7)  18621 (47.3) |
| **Age** |  |
| <65  65-74  75-80  80+ | 22275 (56.5)  6072 (15.4)  3854 (9.8)  7199 (18.3) |
| **Ethnicity** |  |
| White  Asian  Black  Mixed  Other/Unknown | 16703 (42.4)  4711 (12.0)  5018 (12.7)  816 (2.1)  12152 (30.8) |
| **IMD decile (IQR)*** | 4 (3) |
| **≥1 Immunosuppressive condition^** | 5940 (21.1%) |

Date available for *27004 patients and ^28149 patients

**Supplemental Figure S2.** **Number of new infections from the combined Influenza, RSV and SARS-CoV-2 assay by year**

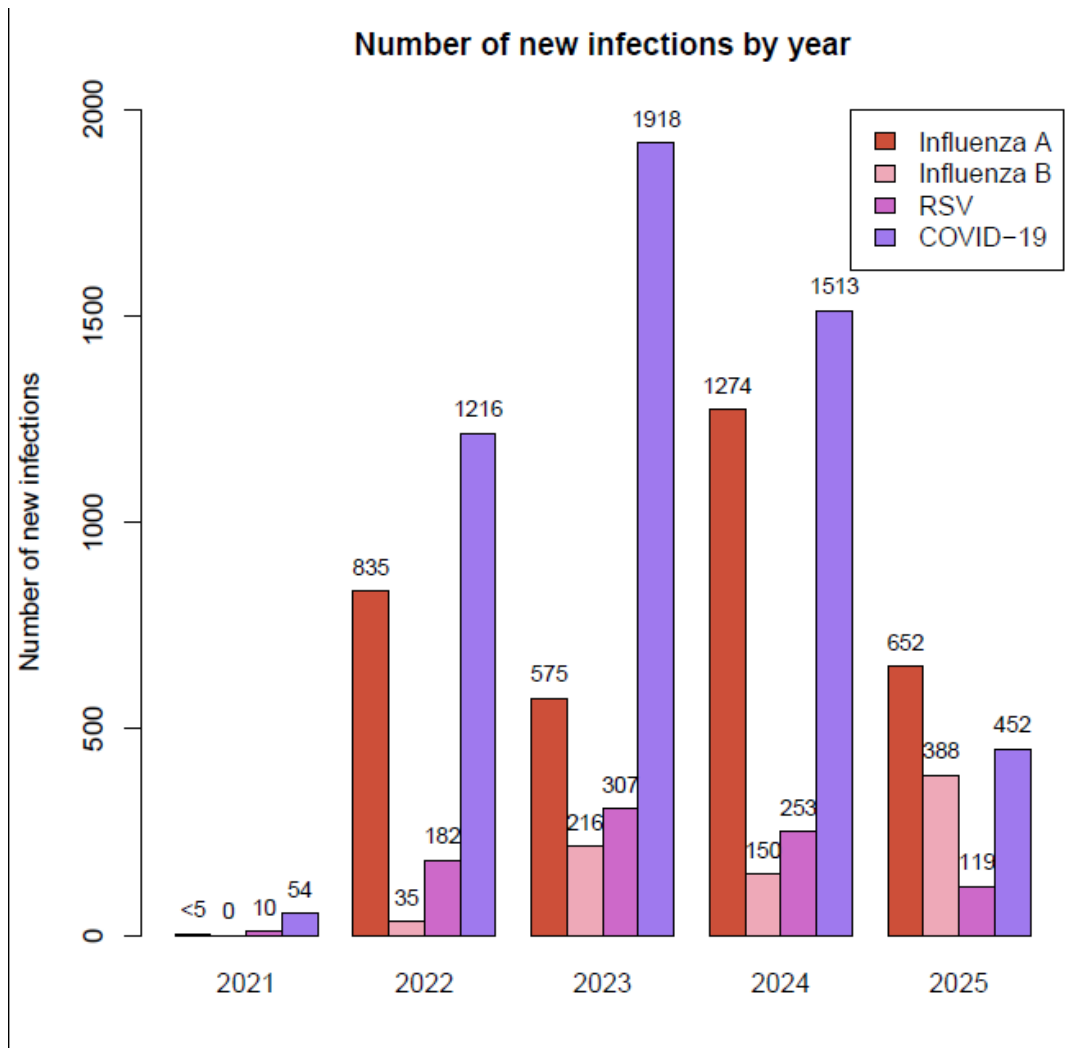

**Supplemental Figure S3. Number of new infections from the combined Influenza, RSV and SARS-CoV-2 assay by season**

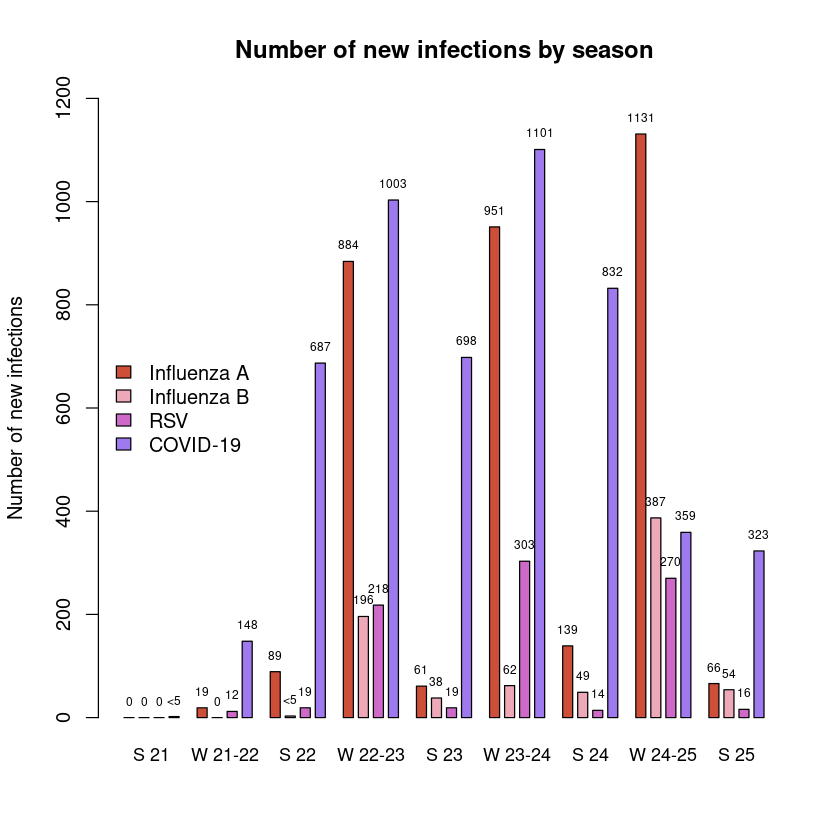

**Supplemental Table S5. Clinical characteristics of 9326 patients testing positive for 9934 Influenza, RSV or SARS-CoV-2 infections**

|  | **Influenza A**  **N= 3310 (%)** | **Influenza B**  **N= 788 (%)** | **RSV**  **N= 864 (%)** | **SARS-CoV2**  **N= 4972 (%)** |
| --- | --- | --- | --- | --- |
| **Sex**  Female  Male | 1878 (56.7)  1429 (43.2) | 484 (61.4)  303 (38.5) | 499 (57.8)  363 (42.0) | 2676 (53.8)  2292 (46.1) |
| **Age**  <65  65-74  75-80  80+ | 2190 (66.2)  401 (12.1)  271 (8.2)  448 (13.5) | 728 (92.4)  24 (3.0)  18 (2.3)  18 (2.3) | 400 (46.3)  162 (18.8)  102 (11.8)  200 (23.1) | 2366 (47.6)  854 (17.2)  595 (12.0)  1157 (23.3) |
| **Ethnicity**  White  Asian  Black  Mixed/Other/Unknown | 1164 (35.2)  474 (14.3)  453 (13.7)  1219 (36.8) | 214 (27.2)  145 (18.4)  112 (14.2)  317 (40.2) | 381 (44.1)  123 (14.2)  104 (12.0)  256 (29.6) | 2157 (43.4)  613 (12.3)  635 (12.8)  1564 (31.5) |
| **IMD decile IQR)*** | 4 (3) | 4 (4) | 4 (3) | 4 (3) |

Data available *IMD=Index of multiple deprivation decile=5996 patients

**Supplemental Table S6. Clinical characteristics of patients with co-morbidity data with severe Influenza A, RSV or SARS-CoV-2 infection requiring intensive care admission and/or dying within 28 days of infection by age and immunosuppressed condition (IC)**

|  | | **Influenza A** | | **RSV** | | **SARS-CoV2** | |
| --- | --- | --- | --- | --- | --- | --- | --- |
| **Severe Infection** | | **N=172(%)** | | **N=89(%)** | | **N=448(%)** | |
|  |  | IC  N=40 (%) | Non-IC  N=132 (%) | IC  N=19 (%) | Non-IC  N=70 (%) | IC  N=131 (%) | Non-IC  N=317 (%) |
| **Age (Years)** | **<65**  **≥65** | 20 (50.0)  20 (50.0) | 62 (47.0)  70 (53.0) | 8 (42.1)  11 (57.9) | 24 (34.3)  46 (65.7) | 52 (39.7)  79 (60.3) | 104 (32.8)  213 (67.2) |
| **ICU Admission** | | **N=108(%)** | | **N=50(%)** | | **N=228(%)** | |
|  |  | IC  N=24 (%) | Non-IC  N=84 (%) | IC  N=9 (%) | Non-IC  N=41(%) | IC  N=67 (%) | Non-IC  N=161 (%) |
| **Age (Years)** | **<65**  **≥65** | 16 (66.7)  8 (33.3) | 57 (67.9)  27 (32.1) | <5* | 23 (56.1)  18 (43.9) | 33 (49.3)  34 (50.7) | 89 (55.3)  72 (44.7) |
| **Death** | | **N=81(%)** | | **N= 50(%)** | | **N=254(%)** | |
|  |  | IC  N=19 (%) | Non-IC  N=62 (%) | IC  N=13 (%) | Non-IC  N=37 (%) | IC  N=74 (%) | Non-IC  N=180 (%) |
| **Age (Years)** | **<65**  **≥65** | 6 (31.6)  13 (68.4) | 10 (16.1)  52 (83.9) | <5* | <5* | 22 (29.7)  52 (70.3) | 23 (12.8)  157 (87.2) |

*Merged due to small number

**Appendix 1**

ICD diagnosis codes for immunosuppressive conditions

| z926 | c817 | c810 | z945 | c944 | c849 |
| --- | --- | --- | --- | --- | --- |
| z948 | c841 | m0699 | k518 | c946 | c861 |
| c900 | m0599 | c884 | c811 | m0596 | c959 |
| z940 | m0590 | c928 | d802 | m074 | d805 |
| y433 | c931 | k508 | d808 | m075 | d807 |
| z857 | m348 | c819 | c865 | m341 | d813 |
| m069 | c831 | c903 | c924 | c824 | m050 |
| z856 | c97x | c822 | d848 | c919 | m0510 |
| c920 | c823 | y431 | m0600 | c966 | m0516 |
| y830 | y434 | m0696 | c820 | l401 | m0589 |
| m059 | d801 | z944 | c848 | m0580 | m0597 |
| t861 | k514 | c829 | c915 | m0693 | m0598 |
| c921 | t868 | z941 | m0739 | c847 | m0603 |
| c833 | m349 | k515 | l403 | c862 | m0606 |
| c911 | c821 | c844 | c827 | c929 | m0680 |
| k509 | m064 | l404 | c852 | c950 | m0689 |
| m350 | c857 | m0694 | c901 | d800 | m0695 |
| k519 | m352 | d849 | d803 | d806 | m0697 |
| l409 | c840 | c835 | m061 | d821 | m0698 |
| c859 | d818 | c809 | z942 | d839 | m071 |
| m321 | c925 | k500 | z947 | d841 | m072 |
| m329 | c917 | c851 | c837 | m052 | m0733 |
| c910 | k512 | k501 | c914 | m0607 | m0736 |
| d591 | m0519 | l400 | d823 | m0769 | m0750 |
| t860 | c902 | c830 | m058 | t862 | m320 |
| k510 | d804 | m0690 | m0594 | z511 | t869 |
| m073 | m068 | k513 | m0609 | z946 | z510 |
| m060 | d838 | c880 | m0730 | c814 | z943 |
| l405 | m328 | l408 | c916 | c839 |  |
| c800 | c838 | m051 | c930 | c846 |  |
